# Comparison of Phase Angle Using Bioelectrical Impedance Analysis According to Device Type and Examination Posture

**DOI:** 10.1101/2022.03.18.22272627

**Authors:** Jihyun Yang, Byung Chul Chun, Jeehyun Kim, Jae-myeong Lee

## Abstract

**Background:** Bioelectrical impedance analysis (BIA) is gaining popularity as a body composition assessment tool for patients. Although it has been studied and validated in different populations, age groups, and clinical settings, including critically ill patients, there are concerns about BIA reproducibility and reliability in different device types and postures. This study aimed to evaluate the reliability of BIA according to different devices, postures, and lead types.

**Methods:** Cross-sectional observational data of 74 healthy volunteers (32 women and 42 men) were collected. We used two types of devices, three types of postures (standing, sitting, and lying), and two lead types (clamp lead and adhesive lead) to measure the whole-body phase angle (phA) at a single frequency of 50 kHz. The measurements were validated using the intraclass correlation coefficient (ICC) and Bland–Altman plot analysis.

**Results:** All phA measurements recorded with two types of devices, three different postures, and two types of leads were equivalent to each other (mean ICC = 0.9932, 95% confidence interval (CI) 0.9905–0.0053). The average mean difference in phA was 0.31 (95% CI 0.16– 0.46). The largest phA value was measured using BWA with an adhesive-type lead in the lying position. There were no differences between standing and sitting positions.

**Conclusion:** This is the first study to show the consistency and reliability of BIA in measuring phA using different devices, lead types, and postures. This could provide the confidence that BIA can be used in various clinical settings.

## Introduction

Body composition measurements can be useful for improving health in the general population, achieving the best performance in athletes, and predicting clinical outcomes and nutritional status in patients (1–3). Bioelectrical impedance analysis (BIA) is a representative method for body composition analysis, using resistance values or impedance resulting from differences in electrical conductivity according to the biological characteristics of the tissues (4). It can be used to evaluate body water composition during treatment planning and monitoring in patients with fluid imbalance. BIA is becoming popular as a body composition assessment tool for patients (1). It has been studied in the general population, patients with malignancy, sarcopenia, obesity, frailty, chronic kidney disease, or cardiovascular disease, as well as in patients in surgical care, and intensive care units (5–15).

However, although BIA has always been a topic of discussion, several limitations have been noted, including the reliability of different algorithms, time of measurement, effect of eating or exercise prior to measurement, ethnicity, sex, and age (15–20). It has also been speculated whether similar/reliable results can be obtained with different devices, measurement methods, postures, and contact locations of the electrode, because of technical limitations (21,22). However, there are no clear data yet.

Various devices have been developed to measure BIA. Inbody^®^ (Inbody Co., Ltd., Korea) used in this study is a product that can quantitatively evaluate water composition in the human body through the measurement of human impedance using multiple frequencies. This study was designed to examine the differences in the BIA method for measuring body composition using different devices, postures, and electrode lead types. We measured the phase angle (phA) at a single frequency of 50 kHz using different methods and analyzed and compared the results.

## Methods

### Study population

This cross-sectional observational study was conducted between May and August 2019. Data were obtained from 74 healthy volunteers, including 32 women and 42 men (Supplementary Information). Written informed consent was obtained from all the participants. Patients who were under 20 years of age, were pregnant, or had a pacemaker insertion before enrollment were excluded. We used the BSM330 (Inbody, Seoul, Korea) to measure the body weight and height of the participants. We compared the different phA measurements. This study was approved by the Korea University Institutional Review Board (IRB No. 2020AN0145), which has been conducted according to the principles expressed in the Declaration of Helsinki.

### BIA measurement protocol/technique

The BIA method was applied as follows: The participants were asked to refrain from consuming any drinks or exercising 4 h before the measurements to minimize disturbance of body fluids. The participants were asked to remain in a standing position for at least 10 min at the beginning of the test. Then, phA was measured for all participants in the standing position using a multi-frequency bioelectrical impedance analyzer (Inbody 970, Inbody, Seoul, Korea) with a grab lead. The device was then changed to BWA 2.0 (Inbody, Seoul, Korea), and phA was measured in the standing position, initially using a clamp lead, followed by an adhesive lead. The participants were asked to switch to a sitting position and were given a 10-min break before the next test. Thereafter, the participants were tested in the sitting position using both the clamp lead and the adhesive lead. Finally, the participants were switched to the lying position, and after a 10-min break, they were tested using the clamp lead followed by the adhesive lead. It took approximately 1 h to complete the entire sequence for each participant. Data on seven measurement methods, including two devices, two lead types, and three postures, were collected. This study was conducted in a single individual.

### Statistical analysis

We compared the results of the seven measurements using the two-way random effects, absolute agreement, and single rater/measurement intraclass correlation coefficient (ICC) method. This implies that our measurement was conducted by a single rater who was selected randomly, and the extent to which the measurements absolutely matched was evaluated (23). ICC values greater than 0.90 imply excellent reliability, whereas values less than 0.5 indicate poor reliability (23). Bland–Altman plots were used to investigate the range of agreement and bias between each measurement (24). Statistical significance was set at p < 0.05 for all analyses. Statistical analyses were conducted using SPSS (version 26.0; IBM, Chicago, IL) and MedCalc 20.008 (MedCalc Software Ltd., Ostend, Belgium).

## Results

The baseline characteristics and average phase angles are presented in Table 1 and Fig. 1, respectively. The data were not normally distributed and are presented as median ± range and mean ± standard deviation.

**Table 1.**
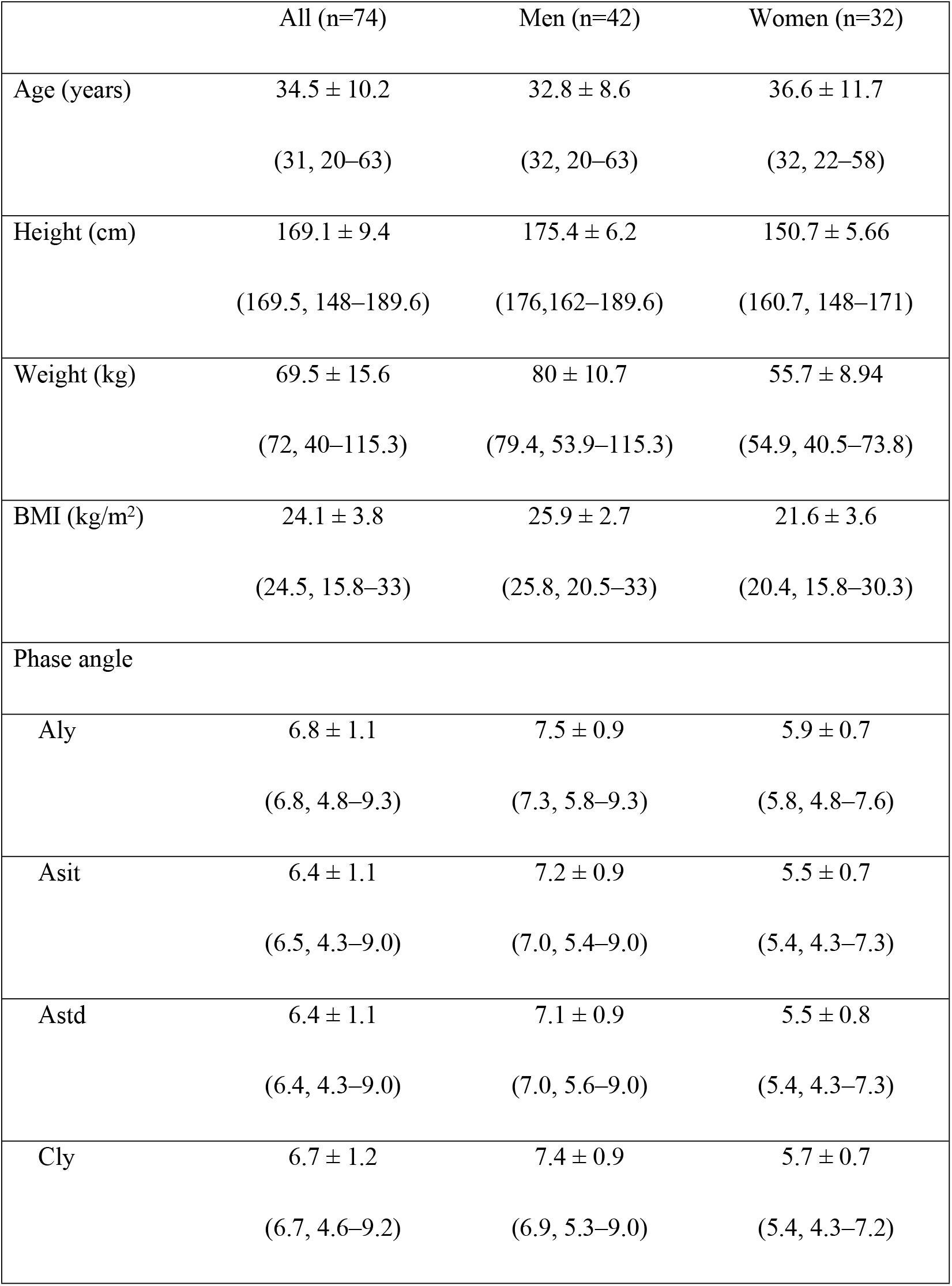
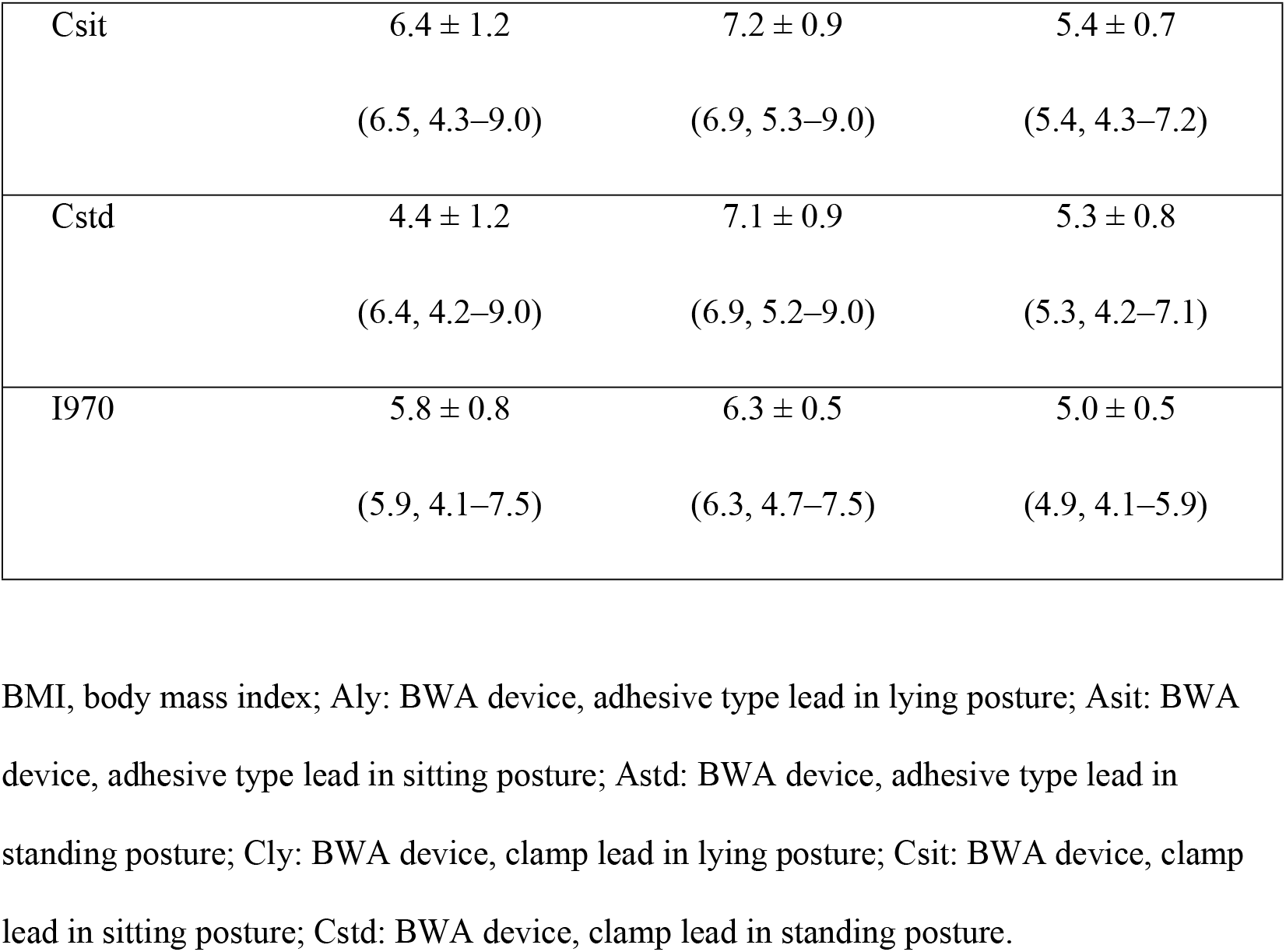
Baseline characteristics of the participants

**Fig. 1.**
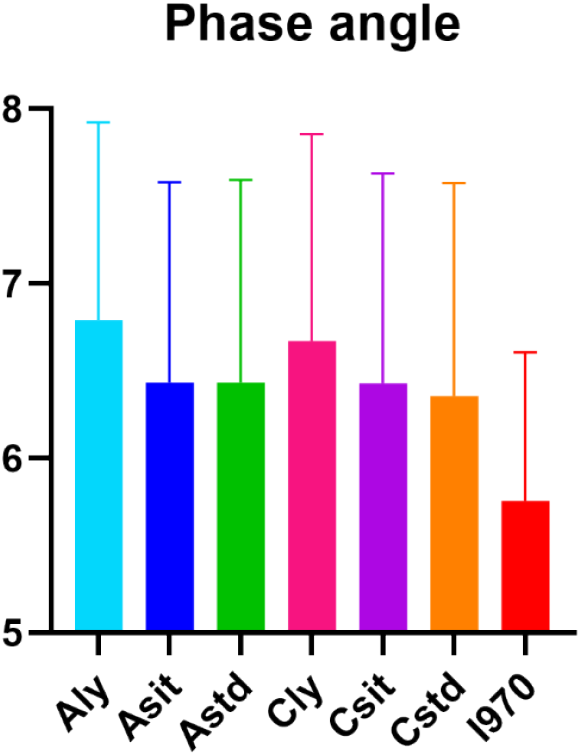
The average of seven different measurements of the 50-kHz phase angle.

### ICC of the phase angle (phA)

The mean ICC of the 50-kHz whole-body phA was 0.9932 (95% confidence interval [CI] 0.9905–0.9953) (Fig. 2).

**Fig. 2.**
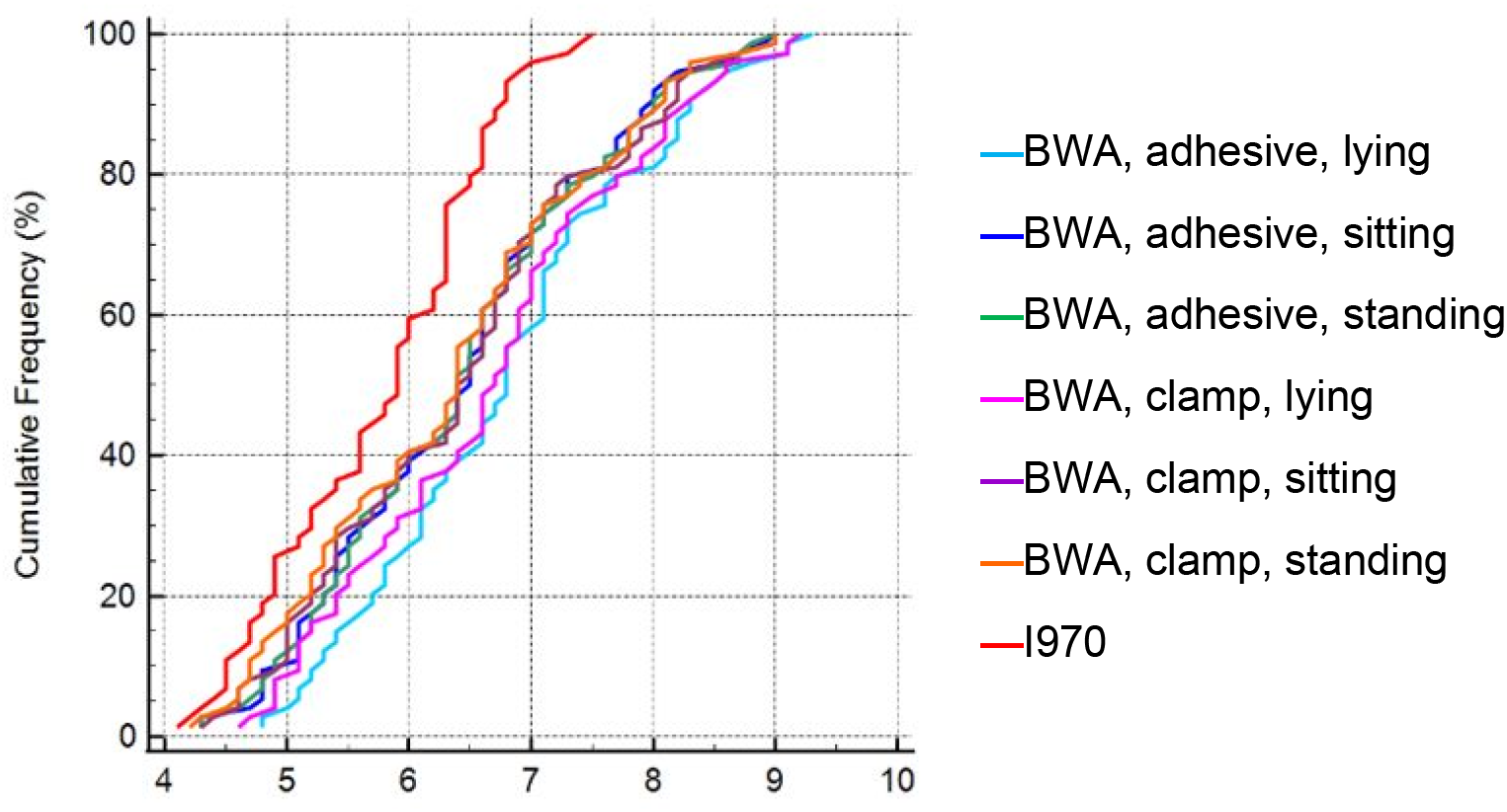
The comparison of intraclass correlation coefficient of 50-kHz whole body phase angle measured using different methods.

### Differences between the seven measurement methods

We further explored the differences in the results using the Bland–Altman plot analysis (Fig. 3, Table 2). The mean difference in the phA was 0.31 (95% CI 0.16–0.46; minimum -0.24, maximum 1.035). No differences in phA were observed between the use of BWA 2.0, adhesive lead in the sitting position, clamp lead in the lying position, and the use of BWA 2.0, adhesive lead in the sitting and standing postures (both mean difference = 0.00, p = 0.95 and 0.99, respectively). The difference between the BWA 2.0 adhesive lead in the lying position and Inbody 970 was the highest (mean difference = 1.04, p < 0.001).

**Table 2.**
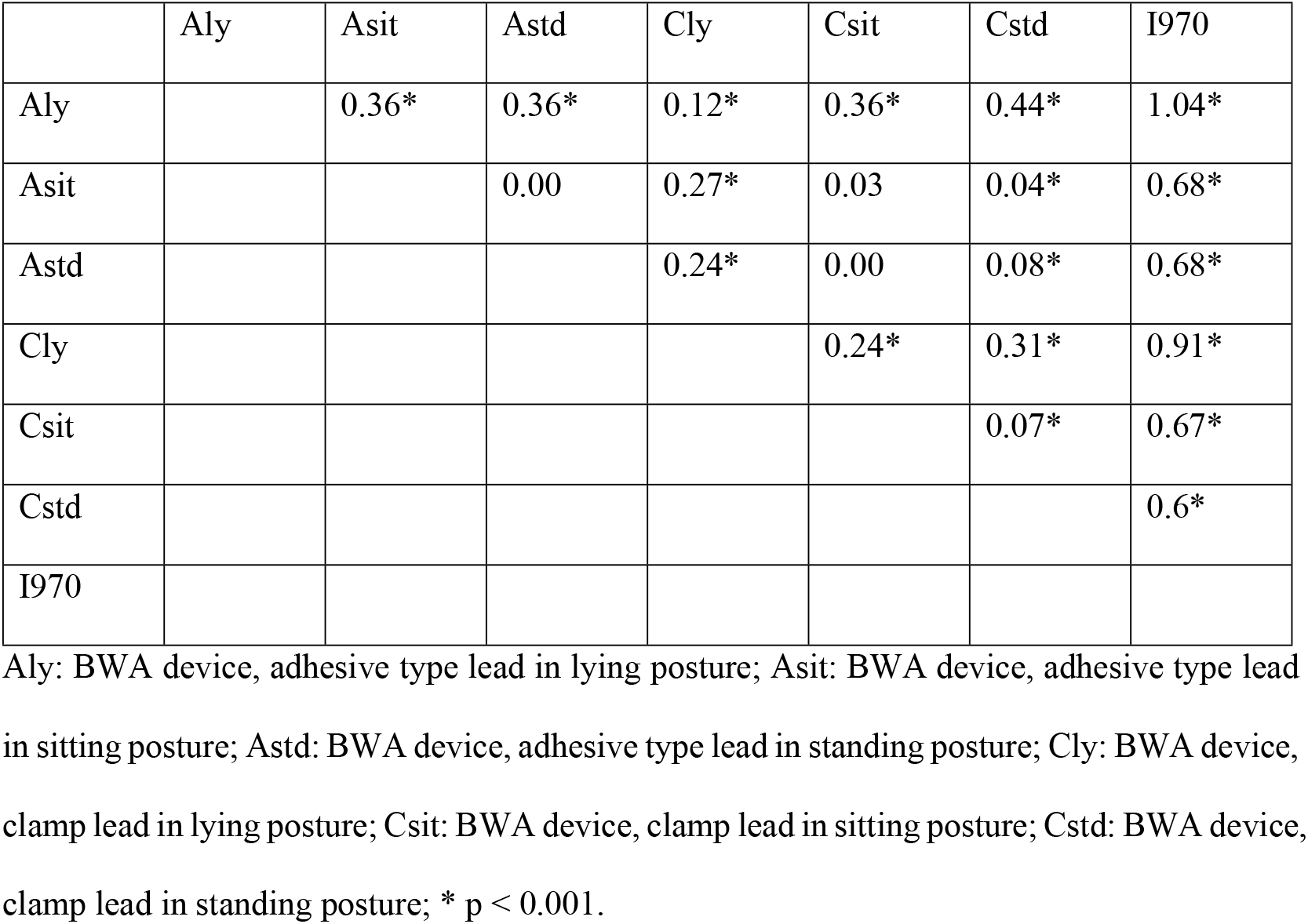
The difference between each 50-kHz 50kHz phase angle measurement according to Bland–Altman B-A plot analysis

|  | Aly | Asit | Astd | Cly | Csit | Cstd | I970 |
| --- | --- | --- | --- | --- | --- | --- | --- |
| Aly |  | 0.36* | 0.36* | 0.12* | 0.36* | 0.44* | 1.04* |
| Asit |  |  | 0.00 | 0.27* | 0.03 | 0.04* | 0.68* |
| Astd |  |  |  | 0.24* | 0.00 | 0.08* | 0.68* |
| Cly |  |  |  |  | 0.24* | 0.31* | 0.91* |
| Csit |  |  |  |  |  | 0.07* | 0.67* |
| Cstd |  |  |  |  |  |  | 0.6* |
| I970 |  |  |  |  |  |  |  |

Aly: BWA device, adhesive type lead in lying posture; Asit: BWA device, adhesive type lead in sitting posture; Astd: BWA device, adhesive type lead in standing posture; Cly: BWA device, clamp lead in lying posture; Csit: BWA device, clamp lead in sitting posture; Cstd: BWA device, clamp lead in standing posture; \* $p < 0.001$ .

**Fig. 3.**
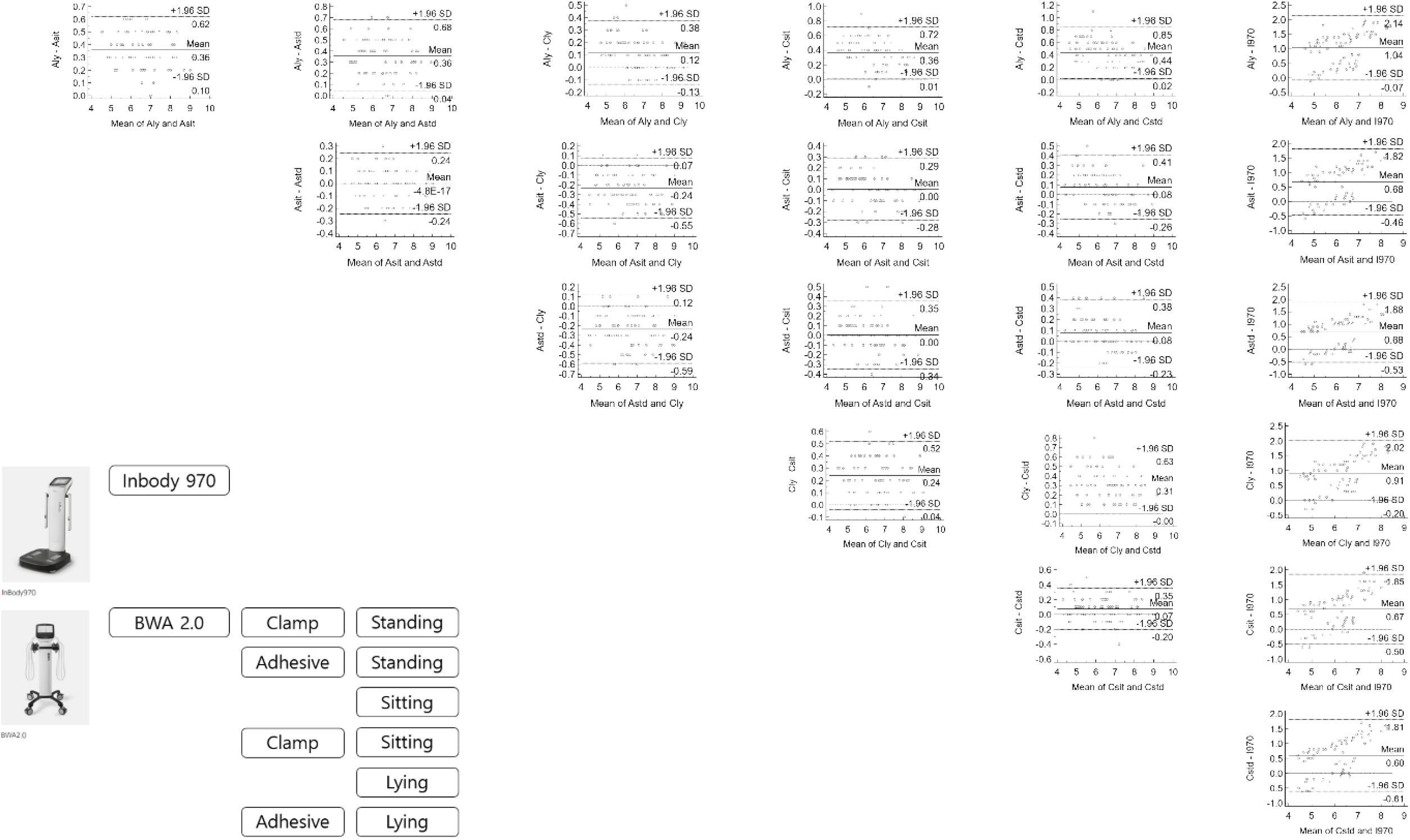
The Bland–Altman plot analysis of different methods of measuring the 50-kHz phase angle.

## Discussion

In this study, we confirmed that the value of phA may not be the same for different devices, postures, and electrodes; however, statistical analyses showed that these values exhibited significant levels of consistency.

BIA measures the volume of fluid and its distribution in the human body using changes in the resistance of the current passing through the fluid with the solute. When this technology was first developed, a single frequency was used to measure the total volume of the human body under the hypothesis that the body has a homogeneous cylindrical structure and current passes through it via electrolytes and water. Since then, further technologies have been developed to measure extracellular and intracellular fluids separately using multiple frequencies, with low-frequency currents not passing through the cell membrane and high-frequency currents passing through it (4). Based on extensive research along with continuous improvements in BIA devices, the European Society for Clinical Nutrition and Metabolism published guidelines for their utilization in clinical practice (20). Although BIA can be useful and appropriately applied in certain populations, particularly in the general population, it has several limitations. This study provides an answer to one of these limitations.

The Bland–Altman plot method only defines the intervals of agreements; it does not indicate whether these limits are acceptable. Acceptable limits must be defined a priori based on clinical necessity, biological considerations, or other goals (24). Using the Bland–Altman plot to compare each parameter, we identified statistically significant differences between the different methods of measuring body composition; however, no clinically significant difference was observed. Previous studies conducted in the general healthy population in Iran and Taiwan reported body phA of 7.32 ± 1.17 and 6.0 ± 0.8, respectively (25,26).

Notably, Inbody 970 showed the smallest value and BWA 2.0, with the adhesive electrode in the lying position, showing the largest value. In addition, the measurement using BWA 2.0, with an adhesive electrode in the standing position, was the same as that with clamping in the sitting position. Because the exact equation is not disclosed, it is difficult to determine the cause of this difference. It is speculated that the adhesive electrode method may detect electricity flow and reflect the characteristics of the body components better than the clamping method, and that the lying position is the most stable.

This is the first study to present consistency in results between different methods of measuring body composition using statistical analysis. The strength of this study is that we directly compared the results of different methods for each subject. However, this study has some limitations. First, we did not compare dual X-ray absorptiometry, which is the gold standard for body composition measurements. Second, the measurements were conducted only in Asian populations. Third, elderly individuals aged >75 years were not included, and the study population comprised relatively middle-aged adults. Fourth, we did not compare all devices; only two representative devices were used.

In conclusion, to the best of our knowledge, this is the first study to show the consistency and reliability of BIA for measuring phA using different devices, lead types, and postures. This study provides information that BIA measured values in various situations can be used interchangeably.

## Data Availability

Data cannot be shared publicly because of IRB regulation.

## Acknowledgments

Not applicable

